# *Streptococcus agalactiae*, a frequent but not so well-known cause of bone and joint infections: a Multicentric observational study 2014-23

**DOI:** 10.64898/2026.03.30.26349534

**Authors:** Simon Jamard, Gwenael Le Moal, Chloe Plouzeau-Jayle, Cedric Arvieux, Sophie Ressier, Raphaël Lecomte, Stéphane Corvec, Séverine Ansart, Claudie Lamoureux, Pierre Abgueguen, Rachel Chenouard, Marie-Frédérique Lartigue, Adrien Lemaignen, the CRIOGO study Group

## Abstract

**Introduction:** *Streptococcus* is the second genus involved in bone and joint infections (BJIs) after *Staphylococcus. Streptococcus agalactiae* is the predominant *Streptococcus* species implicated in BJIs. However, unlike *Staphylococcus*-related BJIs, data on *S. agalactiae* infections remain scarce.

**Methods:** We conducted a retrospective cohort study from the West Region cohort of the CRIOAc registry among six university hospitals including all microbiologically confirmed streptococcal BJI in adults between 2014 and 2023.

**Results:** 1454 patients were included, with a median age of 67 years and 65% male. *S. agalactiae* was the predominant streptococcal species involved 423/1454(29%). The most prevalent comorbidities identified were obesity (378/1454;26%) and diabetes mellitus (343/1454;24%). Prosthetic joint infections (PJIs) were the most common (653/1454;45%), although diabetic foot osteitis was less prevalent overall, it was significantly more associated with *S. agalactiae* infections (48/423;11% *versus* 70/1031;7%, p=0.05). *S. agalactiae* BJIs were more frequently lower-limb infections and chronic infections (240/423;57% *versus* 502/1031;49%, p=0.04). Half of the cohort had a polymicrobial infection and were slightly more frequent with *S. agalactiae* BJIs (235/423;56% *versus* 498/1031;48%, p=0.1). These results were consistent with a sensitivity analysis excluding diabetic foot related osteitis. Logistic regression analysis identified arteriopathy (OR: 4.16; IC95:1.64-11.24, p=0.003), and obesity (OR: 2.57; IC95: 1.41-4.78, p=0.002) as specific risk factors for *S. agalactiae* BJIs.

**Conclusion:** *S. agalactiae* emerges as a prominent and distinct pathogen in complex streptococcal BJIs, with specific risk factors such as arteriopathy, obesity and diabetes mellitus, and more chronic infections.

## Introduction

Bone and joint infections (BJIs) are complex and heterogeneous conditions associated with significant morbidity [1–3] and mortality [4,5]. Their complexity arises from the diversity of infection sites, the chronic nature of some cases, the presence of prosthetic material, the underlying pathophysiological mechanisms, and from the causative microorganisms.

Among these, the *Streptococcus* genus ranks second to *Staphylococcus* as a leading cause of BJIs [6], with a rising incidence [7]. *Streptococcus agalactiae* is the predominant *Streptococcus* species implicated in BJIs [8,9]. However, unlike *Staphylococcus*-related BJIs, clinical and prognostic data on *S. agalactiae* infections remain scarce. Although several comorbidities condition, classically described as risk factors for *S. agalactiae* infection, including diabetes mellitus, obesity, and peripheral arterial disease, are frequently reported, their independent contribution to disease risk remains difficult to quantify. In some cohorts, the prevalence of diabetes in *S. agalactiae* BJIs is reported as high as 54-58% [8,9], yet it drops to less than 20% in others [10–13]. Other factors such as malignancy, cirrhosis, and renal failure are inconsistently reported across studies.

Crucially, the current literature suffers from a lack of direct comparative studies and matched control groups. Most of the available data are derived from retrospective case series or longitudinal surveillance. This makes it impossible to evaluate risk factors directly against a population that is either not infected or infected in a different way. Without such comparisons, it remains unclear whether these comorbidities are specific drivers of streptococcal infections (and more specifically *S. agalactiae* invasion) or merely common characteristics of the aging, comorbid population that typically undergoes arthroplasty

Furthermore, the current literature on the management and prognosis of streptococcal BJIs presents significant contradictions. Historically, these infections were considered responsive to conservative management and associated with favourable outcomes, given the pathogen’s high penicillin susceptibility. However, recent large-scale multicenter studies have challenged this assumption, documenting unexpectedly high failure rates reaching 42% for prosthetic joint infections (PJIs) managed with debridement, antibiotics, and implant retention (DAIR) [14]. Furthermore, *S. agalactiae* has been identified as an independent risk factor for treatment failure in several cohorts, suggesting specific virulence determinants that remain poorly understood compared to other streptococcal species.

As part of the SGBJI project (Streptococcus-Group B/Bone and Joint Infections), aiming to identify clinical and microbiological factors associated with *S. agalactiae* BJIs, we aimed here to describe the epidemiology of streptococcal BJIs with a specific focus on *S. agalactiae* related infections.

## Methods

### Design and data collection population study

We conducted an observational retrospective cohort study from the West Region cohort of the CRIOAc registry [15] (*Centres de référence pour les Infections Ostéo-Articulaires Complexes*, French national network for complex BJIs), gathering six university hospitals. This database contains demographics information, medical history, clinical diagnosis, microbiological documentation, and antimicrobial and surgical therapy. These data were collected during multidisciplinary meeting (MM) gathering at least an infectious disease physician, a microbiologist, and an orthopaedic surgeon. All data were extracted from this database with a quality control of these data from electronic patient records for 29% of the local cohort (420/1454). The concordance rate of the data was greater than 99.5%.

### Study population

To assess specific risk factors associated with *S. agalactiae* BJIs compared to other streptococcal BJIs, we included every adult (>18y/o), with a microbiologically documented streptococcal BJI confirmed in MM: compatible clinical and radiological presentation with streptococcal positive microbiological samples (blood culture, joint fluid and/or surgical sample). The inclusion period covered ten years: from 1^st^ January 2014 to 31^st^ December 2023.

### Clinical definitions

Chronic infections were defined as infections lasting more than four weeks since the first sign of infections. Device related infections were classified as Early, Delayed and Late infections when occurring in the first month, between the first and the twelfth month and after the twelfth month since the device implantation, respectively [16]. Chronic kidney failure was defined as an estimated Glomerular Filtration Rate (CKD-EPI) lesser than 30mL/min/1.73m^2^ and obesity as a Body Mass Index greater than 30kg/m^2^.

### Statistical analysis

All statistical analyses were conducted using R software (version4.5.1, The R Foundation for Statistical Computing, https://www.R-project.org/) and RStudio (v2025.5.1.0). Descriptive statistics were used to summarize baseline characteristics. Continuous variables were compared using Student’s t-test or the Mann-Whitney test, depending on data distribution. Categorical variables were analysed using the χ^2^ test or Fisher’s exact test, as appropriate. To account for multiple comparisons in descriptive analyses, p-values were adjusted using the Benjamini-Hochberg procedure, to control the false discovery rate and limit the risk of type I errors. Statistical significance was defined as an adjusted p-value<0.05. A logistic regression model was employed to identify factors independently associated with *S. agalactiae* BJIs. Variables with a p-value < 0.2 in univariate analysis were considered for inclusion in the multivariate model [17], following assessment for potential interactions. The final model was selected based on minimization of the Akaike Information Criterion (AIC), ensuring optimal balance between model fit and complexity [18]. Missing data were inferior to 5% and were excluded from analysis.

Tables and figures were made with R (v4.5.1) and GraphPad Prism (v8.0.2), and graphical abstract was made with Krita digital painting application v5.2.10 (Krita Foundation).

## Results

Between 2014 and 2023, a total of 18 099 patients were evaluated for BJIs in MM across the six university hospitals affiliated with the West Region CRIOAc network. Among them, 1 646 (9%) were initially identified as possible streptococcal BJI. After exclusion of 192 patients (either no BJI confirmed or microbiological discordance), a final cohort of 1 454 (8%) patients with microbiologically confirmed streptococcal BJIs was retained for analysis, of which 423/1454(29%) were due to *S. agalactiae* (Figure 1).

**Figure 1:**
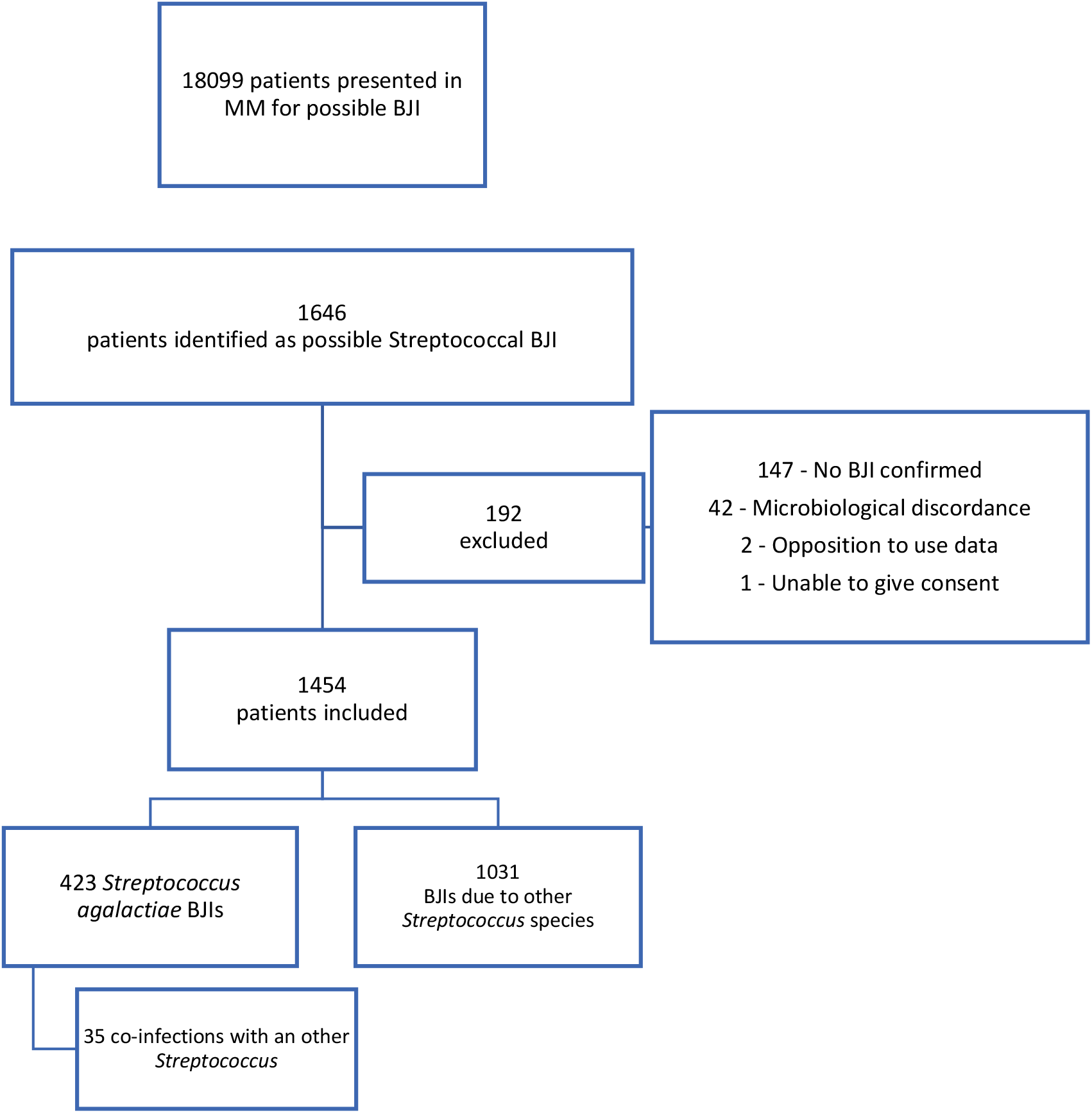
Flow chart. Patients were excluded for “No BJI confirmed” corresponding to patients presented in MM for BJI suspicion without diagnostic confirmation, and for “Microbiological discordance” corresponding to a discordance between the database and electronic patient records or no *Streptococcus* infection confirmed. MM: Multidisciplinary Meeting; BJI: Bone and Joint Infections.

### Population characteristics

The cohort was predominantly male (949/1454;65%) with a median age of 67 years old (IQR: 23) (Table 1). Overall, the population exhibited a moderate burden of comorbidities with an adjusted Charlson score at 3 (IQR:3). The most prevalent comorbidities were obesity (378/1454;26%) and diabetes mellitus (343/1454;24%).

**Table 1:** Baseline characteristics of streptococcal bone and joint infections. A complementary analysis excluding patients with a diabetic foot related osteitis is available in supplementary data. BJI: Bone and joint infections; BH-adj p-value: adjustment of the p-values using the Benjamini-Hochberg procedure.

|  | *S. agalactiae* BJI<br>(n=423) | Other *Streptococcal*<br>BJI (n=1031) | Total<br>(n=1454) | *p-value*<br>BH-adj |
| --- | --- | --- | --- | --- |
| **Population characteristics** |  |  |  |  |
| Sex (Male) | 275 (65%) | 674 (65%) | 949 (65%) | 1 |
| Age Median (IQR) | 66 (22) | 67 (24) | 67 (23) | 0.36 |
| **Past medical history** |  |  |  |  |
| Adjusted Charlson score Median (IQR) | 3 (3) | 3 (3) | 3 (3) | 0.31 |
| Chronic kidney failure *eGFR*<30mL/min | 40 (9%) | 83 (8%) | 123 (8%) | 0.66 |
| Chronic cardiac failure | 56 (13%) | 131 (13%) | 187 (13%) | 0.94 |
| Myocardial infarction | 21 (5%) | 36 (3%) | 57 (4%) | 0.51 |
| Hypertension | 58 (14%) | 119 (12%) | 177 (12%) | 0.51 |
| Arteriopathy | 46 (11%) | 55 (5%) | 101 (7%) | **0.03\*** |
| Stroke | 8 (2%) | 21 (2%) | 29 (2%) | 1 |
| Paraplegia | 36 (9%) | 72 (7%) | 108 (7%) | 0.59 |
| Dementia | 14 (3%) | 30 (3%) | 44 (3%) | 0.93 |
| Obesity *BMI*>30kg/m<sup>2</sup> | 126 (30%) | 252 (24%) | 378 (26%) | 0.17 |
| Malnutrition | 12 (3%) | 18 (2%) | 30 (2%) | 0.51 |
| Chronic pulmonary failure | 22 (5%) | 43 (4%) | 65 (4%) | 0.67 |
| Chronic liver failure | 19 (4%) | 40 (4%) | 59 (4%) | 0.89 |
| Diabetes mellitus | 130 (31%) | 213 (21%) | 343 (24%) | **0.03\*** |
| Neoplasia | 34 (8%) | 57 (6%) | 91 (6%) | 0.29 |
| Hemopathy | 4 (1%) | 12 (1%) | 16 (1%) | 1 |
| Immunosuppression | 36 (9%) | 65 (6%) | 101 (7%) | 0.38 |
| HIV | 1 (0%) | 1 (0%) | 2 (0%) | 1 |
| Inflammatory rheumatism | 28 (7%) | 39 (4%) | 67 (5%) | 0.13 |
| Active smoking habits | 52 (12%)<br>n=397 | 131 (13%)<br>n=986 | 183 (13%)<br>n=1383 | 1 |
| Chronic alcoholism | 39 (9%)<br>n=397 | 72 (7%)<br>n=986 | 111 (8%)<br>n=1383 | 0.36 |

**Table 2:** Bone and joint infections characteristics according to the *Streptococcus* species involved. †Infected sites were counted separately for multiple site infections BJI: Bone and joint infections; BH-adj p-value: adjustment of the p-values using the Benjamini-Hochberg procedure.

|  | *S. agalactiae* BJI<br>(n=423) | Other *Streptococcal*<br>BJI (n=1031) | Total<br>(n=1454) | *p*-value<br>BH-adj |
| --- | --- | --- | --- | --- |
| **Site of infections** |  |  |  |  |
| Number of sites affected |  |  |  | 0.51 |
| 1 | 411 (97%) | 985 (96%) | 1396 (96%) |  |
| 2-3 | 10 (2%) | 42 (4%) | 52 (4%) |  |
| >3 | 2 (0%) | 4 (0%) | 6 (0%) |  |
| Localisation |  |  |  | **0.04\*** |
| Lower limb | 370 (87%) | 824 (80%) | 1194 (82%) |  |
| Upper limb | 24 (6%) | 104 (10%) | 128 (9%) |  |
| Axial | 15 (4%) | 34 (3%) | 49 (3%) |  |
| Cranial | 2 (0%) | 23 (2%) | 25 (2%) |  |
| Multiple | 12 (3%) | 46 (4%) | 58 (4%) |  |
| Arthritis† | 36 (9%) | 127 (12%) | 163 (11%) | 0.21 |
| Osteomyelitis† | 90 (21%) | 192 (19%) | 282 (19%) | 0.51 |
| Of which pressure ulcer associated osteitis† | 37 (9%) | 84 (8%) | 121 (8%) | 0.92 |
| Diabetic foot osteitis† | 48 (11%) | 70 (7%) | 118 (8%) | **0.05\*** |
| Osteosynthesis device† | 85 (20%) | 172 (17%) | 257 (18%) | 0.14 |
| Prosthetic Joint† | 170(40%) | 483 (47%) | 653 (45%) | 0.13 |
| Sort of Prosthetic Joint |  |  |  | 0.70 |
| Hip | 91 (22%) | 255 (25%) | 346 (24%) |  |
| Knee | 73 (17%) | 213 (21%) | 286 (20%) |  |
| Shoulder | 2 (0%) | 7 (1%) | 9 (1%) |  |
| Ankle | 1 (0%) | 7 (1%) | 8 (1%) |  |
| Elbow | 1 (0%) | 1 (0%) | 2 (0%) |  |
| Megaprosthesis (Hip + Knee) | 1 (0%) | 0 (0%) | 1 (0%) |  |
| Time between device implantation and infection | n=221 | n=581 | n=802 | 0.90 |
| Early (<4 weeks) | 35 (16%) | 94 (16%) | 129 (16%) |  |
| Delayed (1-12 months) | 70 (31%) | 166 (29%) | 236 (29%) |  |
| Late (>12 months) | 116 (53%) | 321 (55%) | 437 (55%) |  |
| Median time Month - Median (IQR) | 10 (64) | 17.00 (82) | 15.00 (82) | 0.90 |
| Infection pathway | n=63 | n=219 | n=282 | 0.52 |
| Direct inoculation | 9 (14%) | 41 (19%) | 50 (18%) |  |
| Contiguous infection | 28 (43 %) | 74 (34%) | 102 (36%) |  |
| Hematogenous | 26 (42 %) | 104 (47%) | 130 (46%) |  |
| Chronic infection (> 4 weeks evolution) | 240 (57%) | 502 (49%) | 742 (51%) | **0.04\*** |
| First episode | 257 (61%) | 666 (65%) | 923 (64%) | 0.51 |
| **Infection characteristics:** |  |  |  |  |
| Pain | 332 (78%) | 864 (84%) | 1196 (82%) | 0.29 |
| Erythema | 302 (71%) | 719 (70%) | 1021 (70%) | 0.51 |
| Leakage/fistula | 202 (48%) | 432 (42%) | 634 (44%) | 0.12 |
| Bacteraemia | 70 (17%) | 237 (23%) | 307 (21%) | 0.07 |
| Skin & Soft tissue infection | 55 (13%) | 101 (10%) | 156 (11%) | 0.29 |
| Endocarditis | 7 (2%) | 53 (5%) | 60 (4%) | **0.04\*** |
| Septic shock | 17 (4%) | 53 (5%) | 70 (5%) | 0.67 |
| White Blood Cell count (G/L) Median (IQR) | 11.2 (6.6) | 12.7 (8.1) | 12 (7.4) | 0.23 |
| Protein C Reactive (mg/L) Median (IQR) | 100 (200) | 110 (180) | 110 (190) | 0.92 |
| Fever >38°C | 201 (48%) | 562 (55%) | 763 (52%) | 0.12 |

Compared to patients with BJIs caused by other streptococcal species, those with *S. agalactiae-*related infections exhibited a significantly higher prevalence of metabolic syndrome related conditions including diabetes mellitus (130/423;31% versus 213/1031;21%, p=0.03), arteriopathy (46/423;11% versus 55/1031;5%, p=0.03), and a non-significant trend for obesity (126/423;30% versus 252/1031;24%, p=0.17).

### Clinical presentation

Streptococcal BJIs were predominantly localized to the lower limb and typically involved a single anatomical site. Prosthetic joint infections were the most common type (653/1454;45%) followed by osteomyelitis (282/1454;19%), and infections related to osteosynthesis device (257/1454;18%). Although diabetic foot osteitis was less prevalent overall (118/1454;8%), it was significantly more associated with *S. agalactiae* infections (48/423;11% versus 70/1031;7%, p=0.05).

*S. agalactiae* BJIs were more frequently lower-limb infections and chronic infections (240/423;57% versus 502/1031;49%, p=0.04). These infections were associated with less bacteraemia and significantly less infective endocarditis (7/423;1.7% versus 53/1031;5.1%, p=0.04). The main streptococcus associated with endocarditis was *S. gallolyticus* (14 cases, 38% of all *S. gallolyticus* infections), *S. mitis/oralis* (8 cases, 5% of all *S. mitis/oralis* infections) and *S. sanguinis* (7 cases, 16% of all *S. sanguinis* infections)

### Microbiological profile

*S. agalactiae* was the predominant streptococcal species causing BJIs (423/1454;29%) followed by *S. dysgalactiae* (340/1454;23%), *S. mitis/oralis* (176/1454;12%) and *S. pyogenes* (145/1454;10%) (Figure 2). Over the ten-year study period, the absolute number of streptococcal BJIs presented in MM increased in parallel with the overall number of cases discussed. However, the proportion of streptococcal BJIs relative to total BJI cases remained stable (Figure S1).

**Figure 2:**
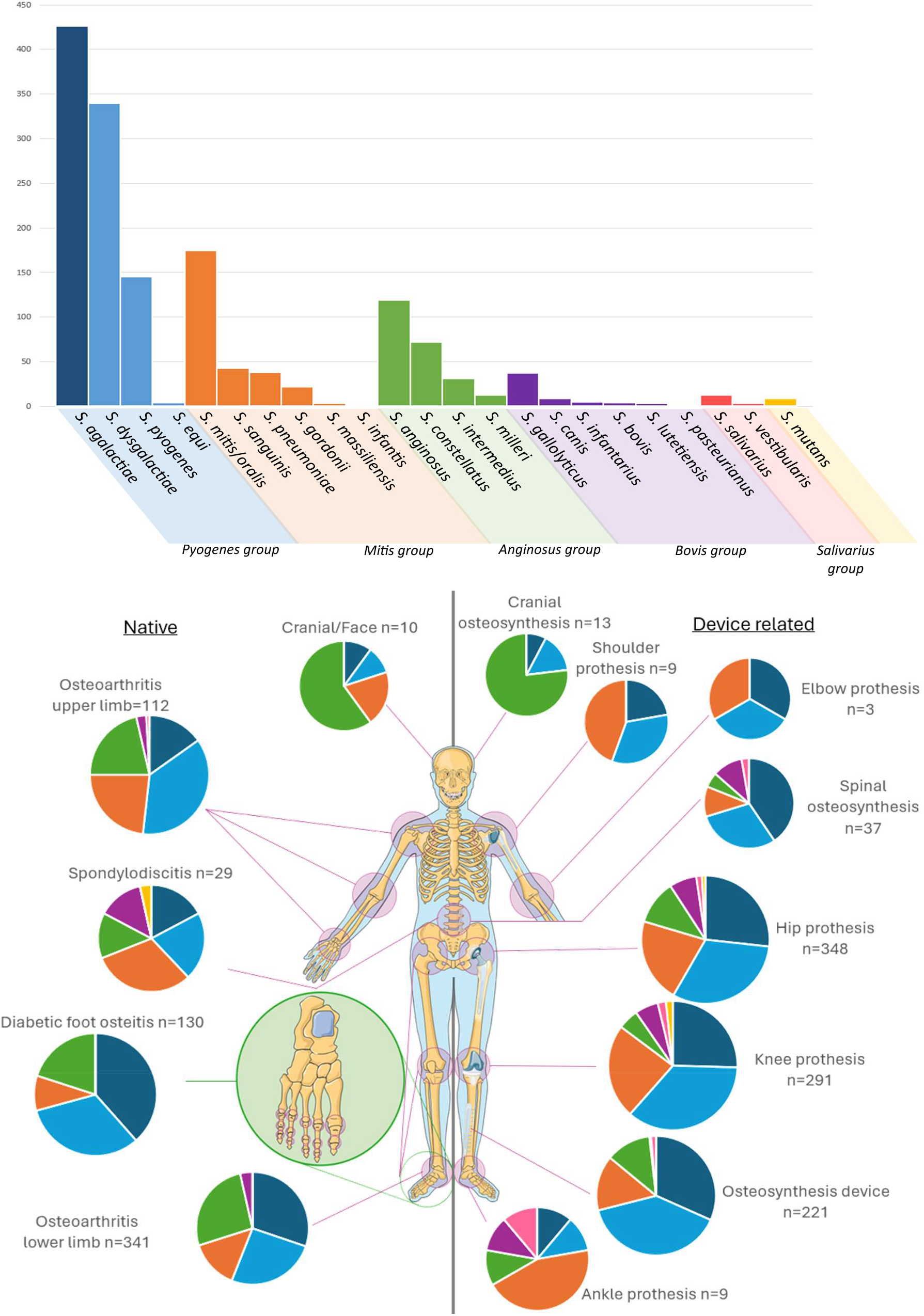
Microbiological repartition of streptococcal BJIs. Repartition of streptococcal species identified in BJIs (A) and ranged between device related or not and localisation, some localisations had more than one *Streptococcus* species. Deep Blue: *S. agalactiae*, Light Blue: other *pyogenes* group, Orange: *mitis* group, Green: *anginosus* group, Purple: *bovis* group, Pink: *salivarius* group, Yellow: *mutans* group. A temporal repartition of the four major strains involved is accessible in supplementary data (Figure S2) *Image adapted from an Image provided by Servier Medical Art (https://smart.servier.com/), licensed under CC BY 4*.*0 (https://creativecommons.org/licenses/by/4.0/)*.

When available antimicrobial susceptibility data are summarized in Table 3. Notably, no penicillin resistance was observed among *S. agalactiae* isolates. In contrast, resistance to penicillin was identified in a limited number of strains from other streptococcal species: four *S. mitis/oralis*, two *S. sanguinis*, one *S. pneumoniae* and one *S. salivarius* strains. Fluoroquinolone remains active against most of streptococcal strains. As anticipated, *S. agalactiae* exhibited a high prevalence of tetracycline resistance, affecting approximately 80% of isolates, consistent with national antimicrobial resistance surveillance data. In contrast, erythromycin resistance was unexpectedly elevated, with 44% of *S. agalactiae* strains resistant compared to national antimicrobial resistance surveillance reported rates of 29.2-34.8% between 2012 and 2024 [19]. We objectivate a growing number *S. agalactiae* isolates exhibiting clindamycin resistance in recent years, suggesting a broader shift toward macrolide/lincosamide resistance within this species (Figure S2).

**Table 3:**
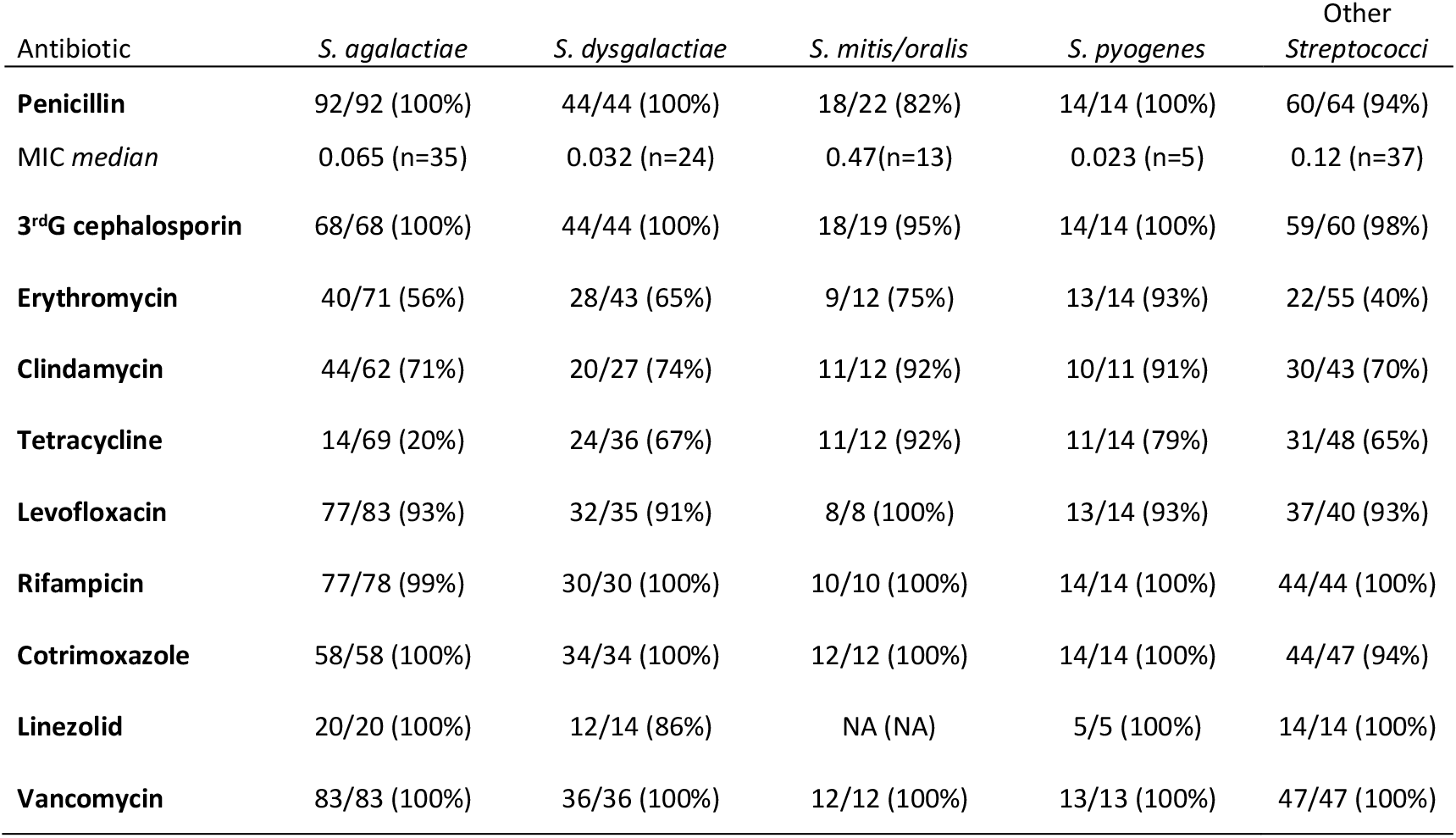
Susceptibilities of streptococcal strains identified in BJIs. Susceptibilities of clinical strains when available expressed as number of susceptible strains on total tested strains and percentage of susceptibility.

| Antibiotic | *S. agalactiae* | *S. dysgalactiae* | *S. mitis/oralis* | *S. pyogenes* | Other<br>*Streptococci* |
| --- | --- | --- | --- | --- | --- |
| **Penicillin** | 92/92 (100%) | 44/44 (100%) | 18/22 (82%) | 14/14 (100%) | 60/64 (94%) |
| MIC median | 0.065 (n=35) | 0.032 (n=24) | 0.47(n=13) | 0.023 (n=5) | 0.12 (n=37) |
| **3<sup>rd</sup>G cephalosporin** | 68/68 (100%) | 44/44 (100%) | 18/19 (95%) | 14/14 (100%) | 59/60 (98%) |
| **Erythromycin** | 40/71 (56%) | 28/43 (65%) | 9/12 (75%) | 13/14 (93%) | 22/55 (40%) |
| **Clindamycin** | 44/62 (71%) | 20/27 (74%) | 11/12 (92%) | 10/11 (91%) | 30/43 (70%) |
| **Tetracycline** | 14/69 (20%) | 24/36 (67%) | 11/12 (92%) | 11/14 (79%) | 31/48 (65%) |
| **Levofloxacin** | 77/83 (93%) | 32/35 (91%) | 8/8 (100%) | 13/14 (93%) | 37/40 (93%) |
| **Rifampicin** | 77/78 (99%) | 30/30 (100%) | 10/10 (100%) | 14/14 (100%) | 44/44 (100%) |
| **Cotrimoxazole** | 58/58 (100%) | 34/34 (100%) | 12/12 (100%) | 14/14 (100%) | 44/47 (94%) |
| **Linezolid** | 20/20 (100%) | 12/14 (86%) | NA (NA) | 5/5 (100%) | 14/14 (100%) |
| **Vancomycin** | 83/83 (100%) | 36/36 (100%) | 12/12 (100%) | 13/13 (100%) | 47/47 (100%) |

Polymicrobial infections were common, observed in half of the cohort (733/1454;50%), and slightly more frequent with *S. agalactiae* BJIs (235/423;56% versus 498/1031;48%, p=0.1) (Figure S3). The most frequently co-isolated pathogens were *S. aureus* (417/733;57%), Gram-negative bacilli (313/733;43%), anaerobic bacteria (214/733;29%), and coagulase-negative staphylococci (125/733;17%). Only Gram-negative bacilli showed a significant association with *S. agalactiae* BJIs (114/235;49% *versus* 199/498;40%, p=0.04).

### Sensitivity analysis

*S. agalactiae* infections were associated with more chronic infections of the lower limb co-infected with Gram-negative bacilli. As these characteristics can also be concordant with diabetic foot related infections (significantly more prevalent in *S. agalactiae* BJIs), we conducted a sensitivity analysis excluding these infections (Table S1). This sensitivity analysis showed a similar significant association with comorbidities (Arteriopathy, diabetes mellitus, obesity), clinical presentation (chronic infection, lower limb infection), and Gram-negative bacilli co-infection, but was not confirmed after Benjamini-Hochberg p-value adjustment.

### Risk factors for S. agalactiae BJIs

Logistic regression analysis identified several comorbidities significantly associated with *S. agalactiae* BJIs. In multivariate model, arteriopathy (OR: 4.16; IC95:1.64-11.24, p=0.003), and obesity (OR: 2.57; IC95: 1.41-4.78, p=0.002) were independently associated with *S. agalactiae* infection (Figure 3). Diabetes mellitus was not identified as a risk factor for these infections (OR: 1.40; IC95:0.75-2.60, p=0.287).

**Figure 3:**
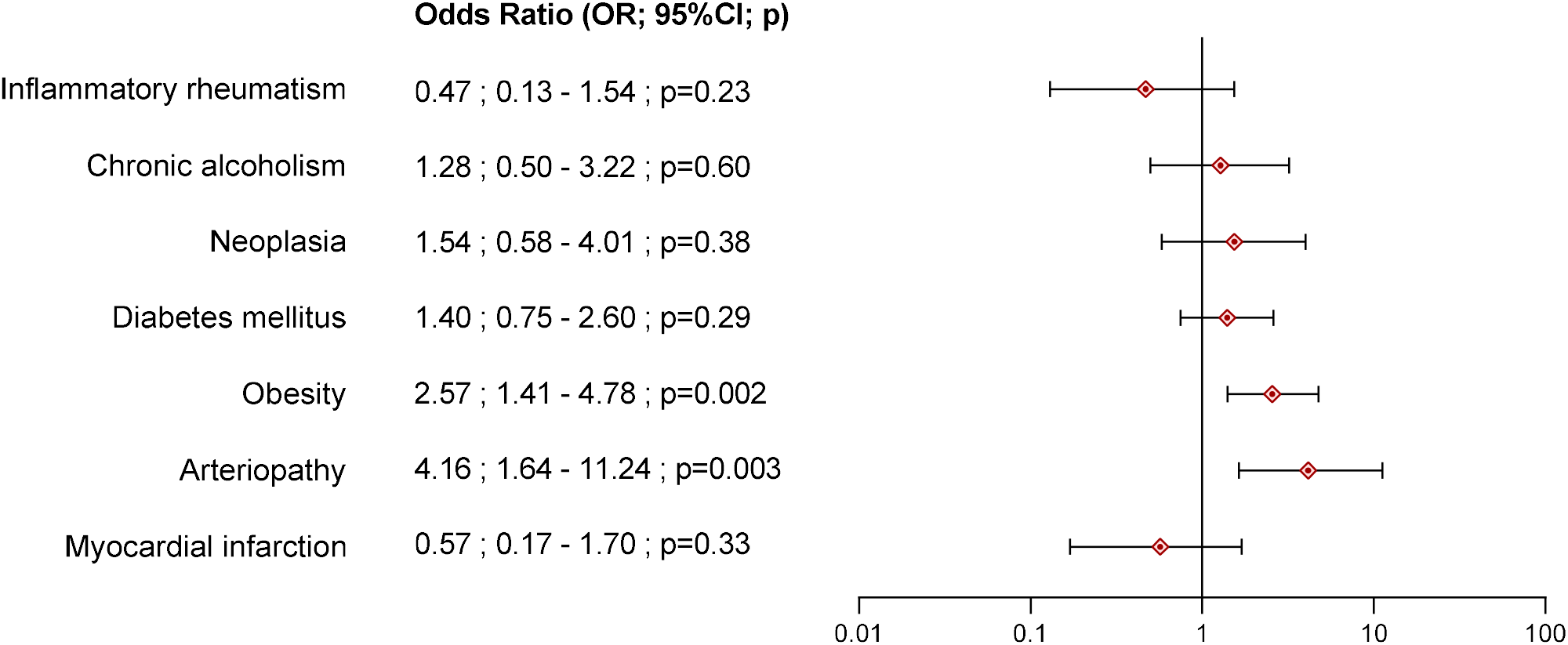
Risk factors associated with *S. agalactiae* BJIs. Odds ratio of comorbidities analysed in multivariate model in logistic regression

## Discussion

This multicentre retrospective study provides a comprehensive overview of streptococcal BJIs across six university hospitals over a ten-year period. Our findings highlight the significant burden of *S. agalactiae* BJIs, which accounted for nearly one-third of all microbiologically confirmed streptococcal BJIs. These infections were notably associated with metabolic comorbidities, chronic clinical presentations and polymicrobial profiles, underscoring their complexity and potential diagnostic challenges.

Among metabolic comorbidities, obesity (defined as a BMI>30kg/m^2^) and arteriopathy emerged as specific factors associated with *S. agalactiae* BJIs. Previous studies have reported similar trends, with high prevalence of diabetes mellitus, obesity, and vascular disease in this population [8,9,20,21]. In our multivariate analysis, diabetes did not remain an independent risk factor, likely due to confounding by a significantly higher incidence of diabetic foot-related osteitis. However, sensitivity analyses excluding diabetic foot-related osteitis, confirmed relevance of this comorbidity. Despite some studies do not shown obesity as risk-factors for *S. agalactiae* BJI [22], several studies identified a high prevalence of obesity in *S. agalactiae* BJI population [20,22,23]. The association between obesity and *S. agalactiae* is well described in pregnancy associated vaginal colonization [24]. Pathophysiology of this association is not definitively identified but could be driven by modification of the gut microbiota [25] and hormonal modification by adipose tissue [26]. In the few studies focusing on this parameter, arteriopathy was also identified with high prevalence coherently to our findings [8,9,21]. This association could be explained by vascular induced micro-wound in skin and gut mucosa promoting hematogenous dissemination of *S. agalactiae*.

Despite the strengths of our large, multicentre cohort and rigorous microbiological confirmation, several limitations must be acknowledged. First, the retrospective design inherently limits data completeness and accuracy. Certain clinical variables, such as hypertension, appeared underreported, likely due to inconsistent documentation in medical records. Second, the absence of follow-up data precluded assessment of patient outcomes, including treatment success, recurrence, and functional recovery. Third, the heterogeneity of infection types - ranging from prosthetic joint infections to diabetic foot osteitis - may introduce variability; however, this diversity reflects real-world clinical practice and enhances the generalizability of our findings. Lastly, our cohort was restricted to complex BJIs managed within the West Region CRIOAc network, potentially excluding cases of native infective arthritis, native vertebral osteomyelitis or diabetic foot infections not requiring multidisciplinary evaluation, which may differ in microbiological and clinical profiles.

Among the complex BJIs in our work, *S. agalactiae* BJIs differ from other streptococcal BJI by their chronicity. They were significantly more chronic infections than other streptococcal BJIs. This could contribute to poorer outcomes of *S. agalactiae* BJIs. While earlier studies suggested favourable prognosis for streptococcal BJIs [13,27], recent evidence indicates more complicated infections, with higher rates of implant-related infections and treatment failure, ranging from 27.1% to 42.1% [8,14,20,28–30]. Zeller *et al*. [31], showed an association between *S. agalactiae* infections and a higher risk of failure HR: 4.88[95% CI: 1.4-17, p=0.012], that could be explain by these more chronic infections and virulence factors that need further investigations.

## Conclusion

In conclusion, *S. agalactiae* emerges as a prominent and distinct pathogen in complex streptococcal BJIs, with specific risk factors such as arteriopathy, obesity and diabetes mellitus, and more chronic infections. Future prospective studies are needed to elucidate prognostic factors and optimize management strategies, particularly in polymorbidity and device-associated infections.

## Data Availability

All data produced in the present study are available upon reasonable request to the authors

## Authors’ contributions

SJ, MFL and AL conceived the protocol. SJ collected the data, ensured the statistical analysis. SJ, MFL and AL wrote the manuscript. All authors participate in Multidisciplinary meeting and care to patients included. All authors reviewed and modified the manuscript. All authors read and approved the final manuscript.

## Acknowledgements

Parts of this work were presented as poster at ESCMIDGlobal 2025 congress and as oral communication at the French National congress of Infectious Disease (*Journées Nationales d’Infectiologie* – JNI) 2025.

We thank all actors of the CRIOGO. Composition of the CRIOGO study group : Adrien Lemaignen, Louis-Romée Le Nail, Marion Lacasse, Frédérique Lartigue, Isabelle Laplaige, Vianney Tuloup, Geoffroy Dubois De Montmarin, Denis Mulleman, Laura Chaufour, Gwénaël Le Moal, Alexandre Losson, Rachel Brault, Chloé Plouzeau-Jayle, Céline Thomas, Diama Ndiaye, Thibault Marty-Diloy, Elisabeth Gervais, Maxime Pichon, Cédric Arvieux, Harold Common, Olivia Berthoud, Vincent Cattoir, Sophie Reissier, Anne Meheut, Marion Baldeyrou, Marie-Clémence Verdier, Raphaël Guillin, Elisabeth Polard, Raphaël Lecomte, Stéphane Corvec, Céline Bourigault, Christophe Nich, Barbara Plantard, David Boutoille, Louise Ruffier D’Epenoux, Reynald Mangeant, François Lataste, Séverine Ansart, Luc Quaesaet, Thomas Williams, Claudie Lamoureux, Jérémy Picard, Anaïs Greves, Pierre Gazeau, Antoine Desseaux, Hervé Le Bars, Pierre Abgueguen, Rachel Chenouard, Florian Ducellier, Emmanuel Hoppe, Frédéric Moal, Amandine Vildy, Hélène Cormier, Hélène Pailhories, Vincent Steiger, Erick Legrand

## Ethics and data protection approval

CRIOAc database is authorized by the French National Data Protection Authority (CNIL-2012/220) and this study has been approved by this authority (registered at University Hospital of Tours under n° 2025_036 and at the public project registration of the Health Data Hub n°22554762). Even in the absence of a requirement for ethical committee approval due to the only use of retrospective routine clinical parameters in accordance with French legislation, we submitted this study for review to institutional ethics committee (Ethics Committee in Human Research, University Hospital of Tours, France), which approved it (approval registration n°2025_015).

## Transparency declaration

None declared.

## References

1 Shichman I, Sobba W, Beaton G, et al. The Effect of Prosthetic Joint Infection on Work Status and Quality of Life: A Multicenter, International Study. J Arthroplasty. 2023;38:2685–2690.e1. doi: 10.1016/j.arth.2023.06.015

2 Gramlich Y, Walter N, Frese J, et al. A Comparison of Health-Related Quality of Life in Patients with Periprosthetic Joint Infection, Patients with Fracture-Related Infections and the General Population-A Multicenter Analysis of 384 Patients from the Section ‘Musculoskeletal Infections’ of the German Society for Orthopaedics and Traumatology. J Clin Med. 2025;14:7649. doi: 10.3390/jcm14217649

3 Frese J, Schwake L, Schulz A-P, et al. Life after bone infection: a retrospective comparison of quality of life in patients with periprosthetic joint infection and fracture-related infections. J Orthop Surg. 2025;20:1012. doi: 10.1186/s13018-025-06427-2

4 Grammatico-Guillon L, Baron S, Gettner S, et al. Bone and joint infections in hospitalized patients in France, 2008: clinical and economic outcomes. doi: 10.1016/j.jhin.2012.04.025

5 Laurent E, Gras G, Druon J, et al. Key features of bone and joint infections following the implementation of reference centers in France. Médecine Mal Infect. 2018;48:256–62. doi: 10.1016/J.MEDMAL.2018.02.004

6 Lemaignen A, Bernard L, Marmor S, et al. Epidemiology of complex bone and joint infections in France using a national registry: The CRIOAc network ✩. J Infect. 2021;82:199–206. doi: 10.1016/j.jinf.2020.12.010

7 Oppegaard O, Skrede S, Mylvaganam H, et al. Temporal trends of β-haemolytic streptococcal osteoarticular infections in western Norway. BMC Infect Dis. 2016;16:535. doi: 10.1186/s12879-016-1874-7

8 Loubet P, Koumar Y, Lechiche C, et al. Clinical features and outcome of Streptococcus agalactiae bone and joint infections over a 6-year period in a French university hospital. PLOS ONE. 2021;16:e0248231. doi: 10.1371/journal.pone.0248231

9 Smith EM, Khan MA, Reingold A, et al. Group B streptococcus infections of soft tissue and bone in California adults, 1995-2012. Epidemiol Infect. 2015;143:3343–50. doi: 10.1017/S0950268815000606

10 Kernéis S, Plainvert C, Barnier J-P, et al. Clinical and microbiological features associated with group B Streptococcus bone and joint infections, France 2004–2014. Eur J Clin Microbiol Infect Dis. 2017;36:1679–84. doi: 10.1007/s10096-017-2983-y

11 Corvec S, Illiaquer M, Touchais S, et al. Clinical Features of Group B Streptococcus Prosthetic Joint Infections and Molecular Characterization of Isolates. J Clin Microbiol. Published Online First: 2011. doi: 10.1128/jcm.00581-10

12 Zeller V, Lavigne M, Leclerc P, et al. Group B streptococcal prosthetic joint infections: a retrospective study of 30 cases. Presse Med. 2009.

13 Meehan AM, Osmon DR, Duffy MCT, et al. Outcome of Penicillin-Susceptible Streptococcal Prosthetic Joint Infection Treated with Debridement and Retention of the Prosthesis. Clin Infect Dis. 2003;36:845–9. doi: 10.1086/368182

14 Lora-Tamayo J, Senneville É, Ribera A, et al. The Not-So-Good Prognosis of Streptococcal Periprosthetic Joint Infection Managed by Implant Retention: The Results of a Large Multicenter Study. Clin Infect Dis. 2017;64:1742–52. doi: 10.1093/cid/cix227

15 Lemaignen A, Grammatico-Guillon L, Astagneau P, et al. Computerized registry as a potential tool for surveillance and management of complex bone and joint infections in France. Bone Jt Res. 2020;9:635–44. doi: 10.1302/2046-3758.910.BJR-2019-0362.R1

16 Société de Pathologie Infectieuse de Langue Française (SPILF). Recommandations de pratique clinique : Infections ostéo-articulaires sur matériel (prothèse, implant, ostéosynthèse). 2009.

17 Heinze G, Dunkler D. Five myths about variable selection. Transpl Int. 2017;30:6–10. doi: 10.1111/tri.12895

18 Dziak JJ, Coffman DL, Lanza ST, et al. Sensitivity and specificity of information criteria. Brief Bioinform. 2020;21:553. doi: 10.1093/BIB/BBZ016

19 Rapport annuel d’activité 2024 - CNR Streptocoques, https://www.cnr-streptocoques.fr/wp-content/uploads/2025/10/RA_2024_vF-sans-annexes-3-4.pdf.

20 Diarra A, Gachet B, Beltrand E, et al. Outcomes in orthopedic device infections due to Streptococcus agalactiae: a retrospective cohort study. BMC Infect Dis. 2024;24:1–5. doi: 10.1186/S12879-024-09175-6/TABLES/2

21 Skoff TH, Farley MM, Petit S, et al. Increasing burden of invasive group B streptococcal disease in nonpregnant adults, 1990-2007. Clin Infect Dis Off Publ Infect Dis Soc Am. 2009;49:85–92. doi: 10.1086/599369

22 Pitts SI, Maruthur NM, Langley GE, et al. Obesity, Diabetes, and the Risk of Invasive Group B Streptococcal Disease in Nonpregnant Adults in the United States. Open Forum Infect Dis. 2018;5:ofy030. doi: 10.1093/ofid/ofy030

23 Sendi P, Christensson B, Uçkay I, et al. Group B streptococcus in prosthetic hip and knee joint-associated infections. J Hosp Infect. 2011;79:64–9. doi: 10.1016/j.jhin.2011.04.022

24 Khalil MR, Hartvigsen CM, Thorsen PB, et al. Maternal age and body mass index as risk factors for rectovaginal colonization with group B streptococci. Int J Gynaecol Obstet Off Organ Int Fed Gynaecol Obstet. 2023;161:303–7. doi: 10.1002/ijgo.14449

25 Musso G, Gambino R, Cassader M. Obesity, Diabetes, and Gut Microbiota. Diabetes Care. 2010;33:2277–84. doi: 10.2337/dc10-0556

26 Klinga K, von Holst T, Runnebaum B. Influence of severe obesity on peripheral hormone concentrations in pre- and postmenopausal women. Eur J Obstet Gynecol Reprod Biol. 1983;15:103–12. doi: 10.1016/0028-2243(83)90178-8

27 Everts RJ, Chambers ST, Murdoch DR, et al. Successful antimicrobial therapy and implant retention for streptococcal infection of prosthetic joints. Anz J Surg. Published Online First: 2004. doi: 10.1111/j.1445-2197.2004.02942.x

28 Andronic O, Achermann Y, Jentzsch T, et al. Factors affecting outcome in the treatment of streptococcal periprosthetic joint infections: results from a single-centre retrospective cohort study. Int Orthop. 2021;45:57–63. doi: 10.1007/s00264-020-04722-7

29 Mahieu R, Dubée V, Seegers V, et al. The prognosis of streptococcal prosthetic bone and joint infections depends on surgical management-A multicenter retrospective study. Int J Infect Dis IJID Off Publ Int Soc Infect Dis. 2019;85:175–81. doi: 10.1016/j.ijid.2019.06.012

30 Fiaux E, Titecat M, Robineau O, et al. Outcome of patients with streptococcal prosthetic joint infections with special reference to rifampicin combinations. BMC Infect Dis. 2016;16:568. doi: 10.1186/s12879-016-1889-0

31 Zeller V, Lavigne M, Biau D, et al. Outcome of group B streptococcal prosthetic hip infections compared to that of other bacterial infections. Jt Bone Spine Rev Rhum. 2009;76:491–6. doi: 10.1016/j.jbspin.2008.11.010

